# Evaluating the impact of keeping indoor dining closed on COVID-19 rates among large US cities: a quasi-experimental design

**DOI:** 10.1101/2021.04.12.21251656

**Authors:** Alina S. Schnake-Mahl, Gabriella O’Leary, Pricila H. Mullachery, Vaishnavi Vaidya, Gabrielle Connor, Heather Rollins, Jennifer Kolker, Ana V. Diez Roux, Usama Bilal

## Abstract

**Objective:** Indoor dining is one of the potential key drivers of COVID-19 transmission. We leverage the heterogeneity in state government preemption of city indoor dining closures, to estimate the impact of keeping indoor dining closed on COVID-19 incidence.

**Methods:** We obtained case rates and city/state re-opening dates from March to October 2020 in 11 U.S. cities. We categorized cities as (treatment) cities that were allowed by the state to reopen but kept indoor dining closed; and (comparison) cities that would have kept indoor dining closed but were preempted by their state and had to reopen indoor dining.

**Results:** Keeping indoor dining closed was associated with a 43% (IRR=0.57, 95% CI 0.46 to 0.69) decline in COVID-19 incidence over 4-weeks compared with cities that reopened indoor dining. These results were consistent after testing alternative modeling strategies.

**Conclusions:** Keeping indoor dining closed contributes to reductions in COVID-19 spread.

**Policy Implications:** Evidence of the relationship between indoor dining and COVID-19 incidence can inform state and local decisions to restrict indoor dining as a tailored strategy to reduce COVID-19 incidence.

## Introduction

The COVID-19 pandemic has directly caused more than 560,000 deaths and 31 million cases through April 2021 in the US alone. Key sources of transmission include crowded and poorly ventilated indoor spaces, including indoor dining areas^1^. Understanding which Non-Pharmaceutical Interventions (NPIs) reduce transmission is crucial for balancing public health and economic and social cost from COVID-19. Previous research suggests that mask mandates^2^, sick leave policies^3^, and shelter in place orders reduce the spread of COVID-19^4^, although the quality of studies concerning the causal evaluation of NPIs is mixed^5^. Many analyses evaluate state-level policies, masking substantial city-and county-level heterogeneity.

During the 2020 summer and fall, states and local governments in the US were left to negotiate closing and re-opening policies without federal regulation or guidance related to activities such as indoor dining. In the early months of the pandemic, most cities and states limited indoor dining, while allowing delivery, take-out, and curb-side pickup services, and sometimes, outdoor dining. As the pandemic progressed some cities and states began reopening indoor dining while others remained closed^6^. Other cities attempted to keep indoor dining closed but were prohibited from doing so, and forced to re-open, by their state.

The federal system in the US delegates much of the authority to protect health to states, under the police powers, and states can then further delegate authority to local governments^7^. The authority to prohibit or limit a lower level of government’s power to enact legislation, is a legislative doctrine known as government preemption^7^. States have preempted local government on laws including minimum wage, bans on removing confederate statues, anti-discrimination polices, and anti-gun legislation^7^. Recently, preemption laws have been adopted by predominantly white and conservative legislatures to limit progressive policies in democratic and majority non-white cities^8^. For example, some states have preempted cities from keeping indoor dining closed, forcing cities to re-open indoor dining during the COVID-19 pandemic^9^.

State preemption of city law around indoor dining created substantial heterogeneity between cities in the scope of local governments’ authority to regulate indoor dining and provides an opportunity to test the association between keeping indoor dining closed and COVID-19 rates. The objective of this study is to examine the association between keeping indoor dining closed and COVID-19 rates, leveraging the variation in local government power to enact these orders. This variation creates a counterfactual contrast that we exploit, by comparing cities that kept indoor dining closed to cities that were forced re-open indoor dining by their respective state governments.

## Methods

### Study Setting

The study population includes cities that are members of the Big Cities Health Coalition (BCHC), an organization comprised of America’s largest metropolitan health departments. We collected information on indoor dining re-opening policies from searching and reviewing city and state orders available on state and city websites.

Following a policy trial emulation framework^10^ we define units of analysis and exposure, causal contrasts, outcomes and time-zero. **Table 1 and Figure 1** contain details on these design specifications, and a summary follows. The main unit of analysis were cities that explicitly expressed an intention to keep indoor dining closed even after their respective states allowed them to reopen indoor dining. Cities were then divided into treatment and comparison groups, according to whether their respective state governments actually allowed them to continue keeping indoor dining closed. The <u>treatment</u> group includes cities that kept indoor dining closed, while the state allowed indoor dining to reopen. The <u>comparison</u> group includes cities that intended to keep indoor dining closed but were preempted from doing so by the state and so reopened indoor dining. By identifying comparison cities that “would have” remained closed absent the preemption, we provide a stronger counterfactual comparison group than comparing all geographies that remained closed to those that re-opened, or studies that compare policy adoption over time^11^, as selection into the treatment group may be biased by other city characteristics.

**Table 1:** Treatment and Comparison Group Definitions.

|  | **Treatment** | **Comparison** |
| --- | --- | --- |
| **Definition** | Cities allowed to re-open by the state, but that <u>stayed closed</u> | Cities that would have kept indoor dining closed but were preempted by their state and <u>re-opened indoor dining</u> |
| **Post-Period Start (time zero)** | Date state <u>allowed</u> the city to re-open, but city stayed closed | Date the city/state <u>re-opened</u> |
| **Included Cities** | Milwaukee, Indianapolis, Philadelphia, San Francisco | Atlanta, Austin, Charleston, Dallas, Houston, Phoenix, San Antonio |
| **Number (n)** | 4 | 7 |
| **Weeks Indoor Dining Closed (before “time zero”)** | 12.9 (7 – 24) | 5.6 (5 - 6) |
| **Incidence rate at “time zero” (per 100,000)** | 10.7 (7.6 - 15.4) | 3.8 (1.3 - 6.9) |
| **Population Size (millions)\*** | 0.96 (0.88 - 1.58) | 2.00 (0.41 - 4.71) |
| **% aged <18\*** | 22.7 (13.4 - 24.5) | 23.5 (19.6-26.4) |
| **% aged >=18-64\*** | 63.5 (62.2 - 70.5) | 63.1 (61.0- 68.7) |
| **% aged >65\*** | 14.0 (12.9 - 16.0) | 12.0 (10.2- 16.8) |
| **% living in poverty \*** | 15.8 (9.5 - 23.3) | 13.5 (10.9 – 15.2) |
| **% college educated\*** | 31.4 (31.0 - 59.2) | 33.5 (28.1 - 57.5) |
| **% Female** | 51.7 (49.2 - 52.7) | 50.6 (49.5 - 51.7) |
| **% overcrowded (>1/room)** | 2.7(1.4 - 7.3) | 4.2(1.5 - 6.4) |
| **Race/Ethnicity** |  |  |
| **% Black (non-Hispanic)** | 27.0(5.2 – 40.1) | 18.54(4.4 - 43.8) |
| **% Hispanic/Latino** | 15.2 (10.9 - 15.6) | 33.6(5.3, 60.68) |
| **% Non-Hispanic White** | 40.1(34.2 – 53.8) | 39.3 (27 – 69.6) |
| **% service workers** | 8.7(8.2-9.5) | 8.3(6.5-10.9) |
| **% foreign-born** | 11.7(9.4 - 33.7) | 14.9(6.7 - 26.3) |
| **%using public transit** | 15.2(1.4 - 36.3) | 2.5(1.5 - 7.0) |
| **% voted for Biden in 2020** | 78.6(63.7 - 85.3) | 58.3(50.3 - 72.6) |
| **# of cities w/ democratic mayors** | 4 (100%) | 6 (86%) |
| **% always or frequent mask wearing** | 84.7(76.1 – 93.8) | 89.2(84.3 – 95.3) |

\* numbers indicated are Medians (Min-Max).

**Figure 1:**
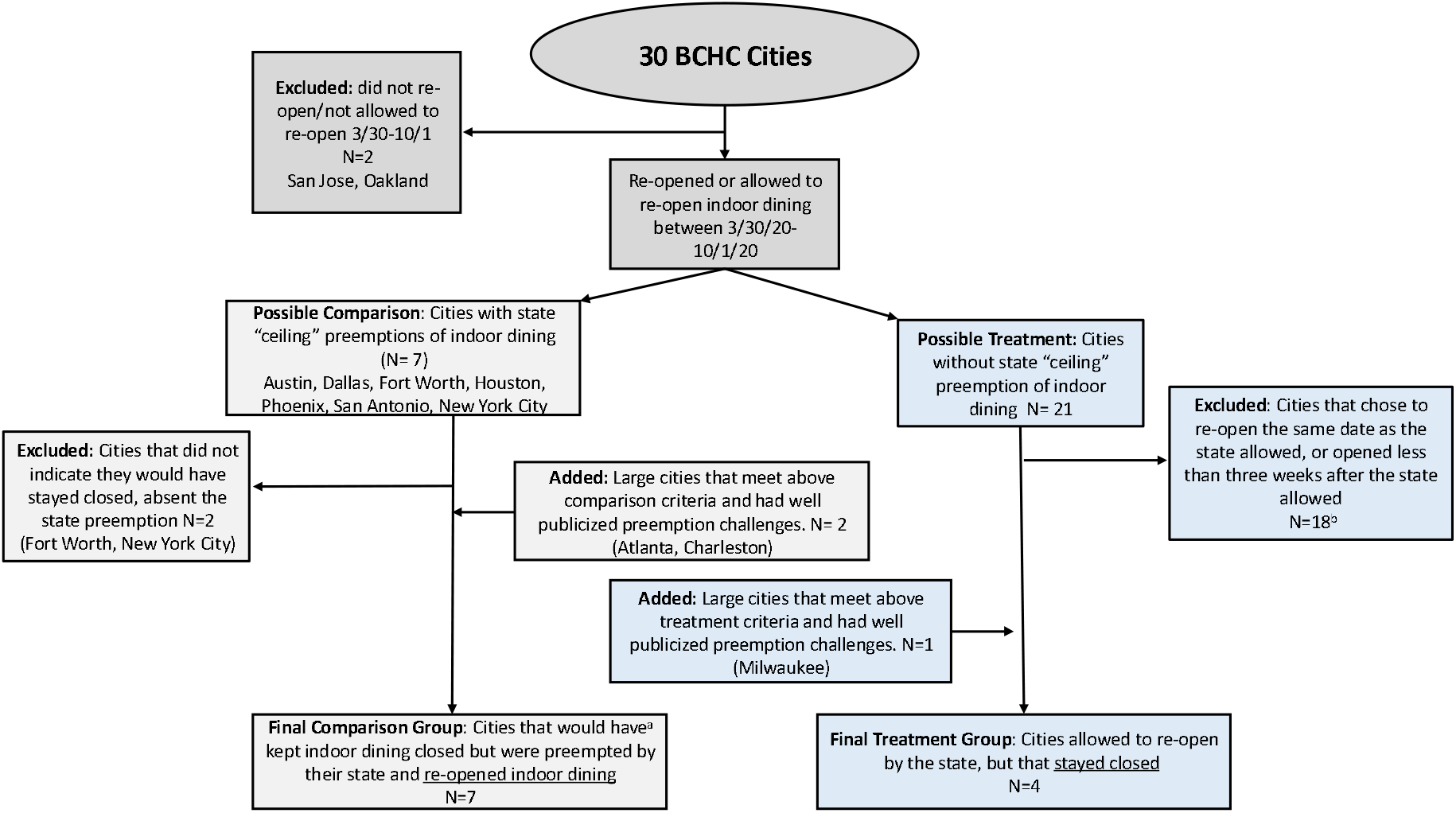
Treatment and Comparison group selection flowchart. ^a^See Appendix B1 for evidence that comparison cities meet the counterfactual condition that they “would have stayed closed” absent the state preemption.

**Figure 1** shows a flowchart of treatment and comparison group selection. We examined the spring/summer period of re-openings after initial stay-at-home orders, and so limited the sample of cities to those that re-opened or were allowed to re-open between March 30, 2020 and October 1, 2020. We excluded cities that did not demonstrate intention to stay closed and then divided cities into treatment and comparison groups. Appendix B provides evidence that comparison cities would have stayed closed absent the preemption order, along with evidence on treatment decisions to remain closed and details on subsequent re-openings in treatment cities. We also excluded treatment cities that re-opened dining less than 3-weeks after the state allowed them to re-open, to ensure a sufficient period of continued closure to impact COVID-19 rates. To increase the number of cities, we included cities that are not in the BCHC but that are among the largest cities in the country, and otherwise met inclusion criteria (opened or allowed to re-open 3/2/20-10/1/20, expressed desire to re-open, and if in treatment, re-opened more than 3-weeks after allowed).

Preemption also helped us to identify “time-zero”^12^. For treatment cities, time-zero is the date the city kept indoor dining closed when the state allowed re-opening, and for comparison cities, this is the date the state mandated (via preemption) the city re-open indoor dining, or when the city, “would have” stayed closed (see Appendix Table A2 for city and state dates). Incorrectly identifying the time when the comparison group “could have” enacted the policy can bias estimates by inappropriately conditioning on or selecting on post-treatment variables, and the preemption allows for easy identification of time-zero in our comparison group^10^. To isolate the immediate impacts of keeping dining closed we limited the study period to 10 weeks total: the 4 weeks before and 6 weeks after “time zero”. Throughout the article, “cities” refers to the city itself, or the county in which the majority of the city resides, depending on data availability (see Appendix Table A2).

### Outcome

We employ city-level (or county equivalent) daily COVID-19 confirmed case and death counts from the BCHC COVID-19 Health Inequities in Cities dashboard^13^, and data from the Center for Systems Science and Engineering (CSSE) at Johns Hopkins University^14^ for cities not available on the BCHC dashboard. To ensure comparability of treatment and comparison cities, we calculated city descriptive statistics, and population denominators for daily city or county level case and death rates, using the 1-year 2019 American Community Survey (ACS)^15^, New York Times county-level mask survey ^16^, and Politico 2020 county-level election results ^17^.

### Covariates

We controlled for other time-varying city-level NPIs, specifically: stay-at-home orders, mask mandates, and eviction moratoriums, derived from multiple policy trackers^18,19^ and review of state and city orders. We did not include starts of stay-at-home orders, school, or religious gathering reopening covariates, as no cities re-started stay-at-home orders and no schools reopened during the study period, and no treatment city restricted religious gatherings as every city was in a state that designated religious gatherings as essential services. We included the NPI variables as binary time-varying indicators (see Appendix A).

### Statistical Analysis

We employed a quasi-experimental approach with a difference-in-difference (DiD) analysis^20^. Cities enacting different indoor dining policies over time allowed us to compare changes in COVID-19 cases before and after the potential re-opening date (1^st^ difference), to changes in COVID-19 cases in areas that did not and did re-open (2^nd^ difference). We assume that the outcome for the comparison group reflects the change that would have been observed in the treatment group had the treatment not occurred^21^. Our study design and analysis method controls for unmeasured time-fixed city confounders, time-varying unmeasured confounders that are similar across cities, and time-varying city-level measured confounders (other NPIs).

We plotted average weekly case rates and timing of state and city indoor dining reopenings for treatment and comparison cities, to visually inspect the relationship between reopenings and case rates. We then used visual examination to test whether cities that kept indoor dining closed had differential pre-policy trends in COVID-19 case and death rates. We measured our treatment as a binary indicator for open(=0)/closed(=1), with open defined by any level of reopening indoor dining, including with limitations (e.g., occupancy limits, special hygiene measures, etc.). We included two-week lags after the “time-zero” to allow for time between exposure and incubation period, testing lags, and reporting lags.

We ran negative binomial regression models, with city-day as the unit of analysis, city population model as an offset, and robust standard errors clustered at the city level. The model specification is available in Appendix C. Using our main model, we calculated the average daily cases prevented by keeping dining closed in the treatment cities, and then estimated the averge marginal effect, calculated as the count of cases that would have been averted in all 11 cities, had no city reopened over the four week period.

We conducted six categories of sensitivity analyses to test for robustness of our model to alternative specifications. Details on the sensitivity analyses are available in Appendix D. In summary, we: (1) extended the duration of follow-up to 12 weeks and varied the pre-specified 2-week lag, (2) excluded various cities, (3) included calendar fixed-effects, (4) used the dates treatment cities actually re-opened indoor dining rather than the dates the state allowed the treatment cities to re-open, (5) used an event study model, an alternative specification of the DiD model, and (6) repeated the analysis using death rates instead of case rates.

We conducted analyses in R 4.0.2 and STATA 15.1.

## Results

We included 11 cities in our analysis, 4 treatment and 7 comparison, across 8 states. The 11 cities contributed a total of 781 city-day observations and accounted for a total of 88,589 cases in the 10-week study period (4 weeks before and 6 weeks after “time-zero”). **Table 1** displays descriptive statistics on treatment and comparison cities: groups were comparable in terms of population size, age distributions, socioeconomic conditions (poverty rates and education levels), percent non-Hispanic white, foreign born, housing crowding, service workers, masking levels, and political leaning. Percentage Hispanic was larger, and percentage non-Hispanic Black smaller, in comparison cities, and public transit use was more common in treatment cities. Indoor dining was closed for an average of 12.8 and 5.6 weeks before cities were allowed to re-open dining (in treatment cities) and before having to open (in comparison cities), respectively. Treatment cities remained closed for a mean of 5.1 weeks after they were allowed to re-open. **Appendix Figure A1** shows the pattern of COVID-19 cases and timing of indoor dining closing and re-opening from April to October; for treatment cities, the figure shows the date states allowed re-opening and the date the city eventually re-opened. In cities that reopened we saw an increase in cases approximately two weeks after indoor dining re-opened, while case rates decreased or continued to decrease in cities that maintained indoor dining closures.

On average, treatment and comparison cities had 11 and 4 daily cases per 100,000, respectively, before reopening indoor dining or keeping indoor dining closed (**Table 1**). But, as shown in **Figure 2** pre-treatment case rate trends were approximately parallel. Results from the models show a strong association between COVID-19 rates and keeping indoor dining closed. Keeping indoor dining closed reduced COVID-19 incidence by 43% (IRR=0.57, 95% CI 0.46 to 0.69) compared with cities that reopened indoor dining, in the fully adjusted model (**Table 2**).

**Table 2:**
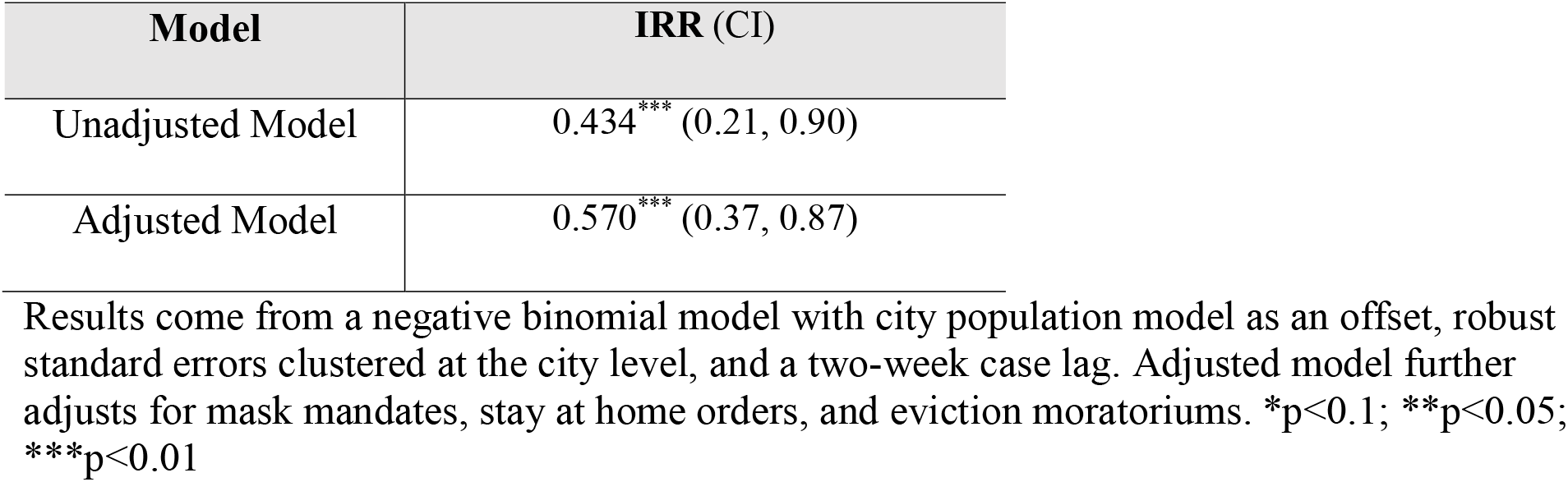
Unadjusted and Adjusted COVID-19 Incidence Rate Ratio (IRR)

| Model | IRR (CI) |
| --- | --- |
| Unadjusted Model | 0.434*** (0.21, 0.90) |
| Adjusted Model | 0.570*** (0.37, 0.87) |

Results come from a negative binomial model with city population model as an offset, robust standard errors clustered at the city level, and a two-week case lag. Adjusted model further adjusts for mask mandates, stay at home orders, and eviction moratoriums. \*p<0.1; \*\*p<0.05; \*\*\*p<0.01

**Figure 2:**
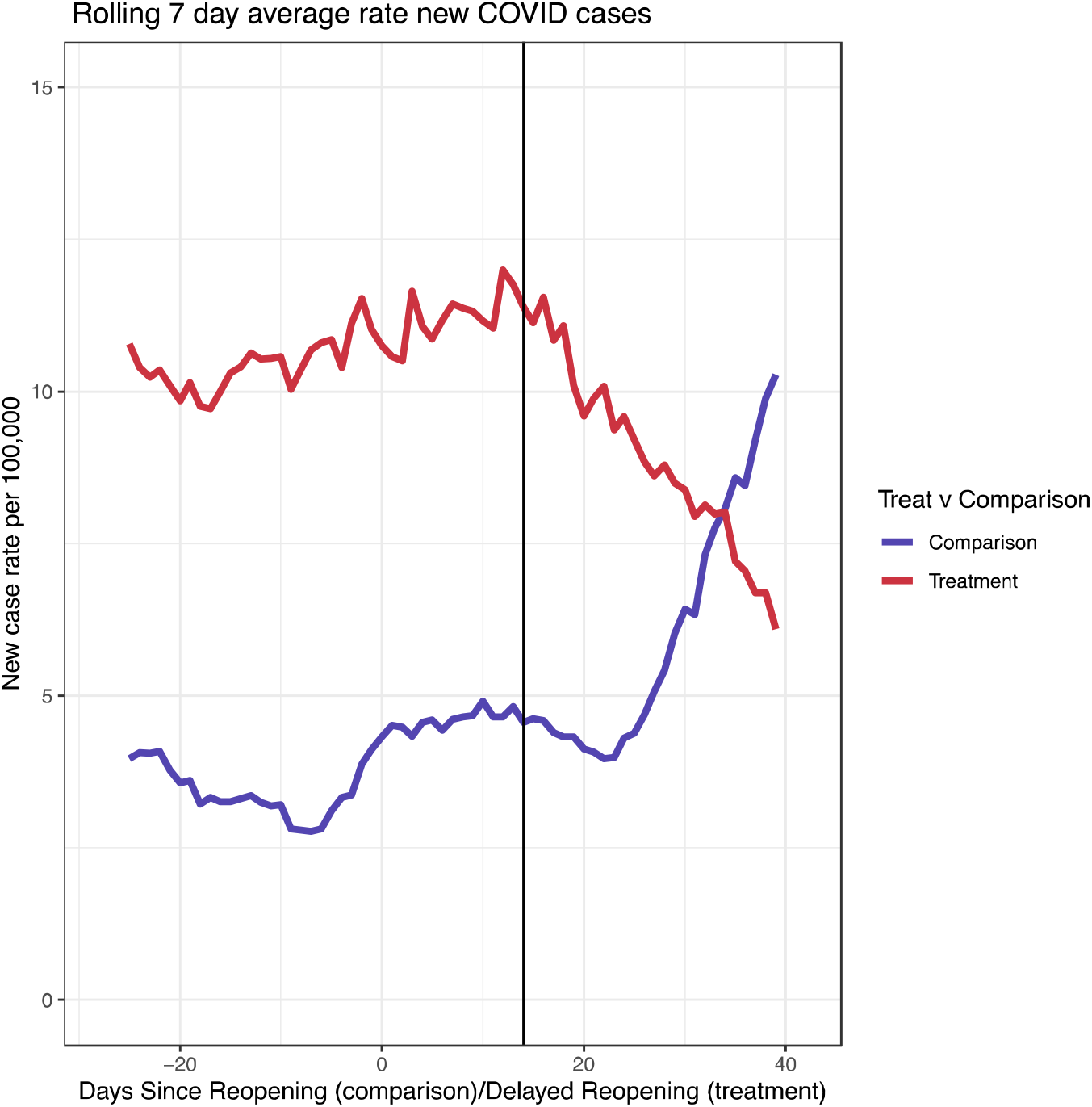
Trends in Case Rates Comparing Treatment and Comparison Groups. Vertical black line represents a 2-week lag after re-opening (comparison cities) or the date the state allowed the city to re-open but the city stayed closed (treatment cities). Comparison cities include Atlanta, Austin, Charleston, Dallas, Houston, Phoenix, and San Antonio; treatment cities include Indianapolis, Milwaukee, Philadelphia, and San Francisco.

Keeping indoor dining closed averted 91 (95% Confidence Interval [CI]: −168, −15) daily cases in the average city. Had all cities kept indoor dining closed (and holding all other NPIs at their actual levels), we estimate that approximately 27,914 (CI: −51253, −4575), or 133 per 100,000, cases would have been averted over the four-week period. These estimates should be viewed cautiously and as general approximations.

### Sensitivity Analyses

The changes to study period, lags, city selection, and inclusion of calendar week fixed effects showed no changes to our inferences (**Appendix Table D2**). Extending the duration of the follow-up period to 12-weeks suggested a larger association, as would be expected given exponential growth. The event model specification reinforces the finding that cases declined in cities that kept indoor dining closed, and that re-opening indoor dining increased cases. In the event study model (**Appendix Table D3 and Appendix Figure D4**), COVID-19 incidence was significantly higher in cities that reopened indoor dining, starting 5 weeks after re-opening, with 3.52 times the incidence compared to the week before re-opening (CI: 1.16, 10.69). The magnitude of these differences increased over the follow-up period; 8 weeks after re-opening indoor dining, cities had 10.85 times higher incidence (CI: 3.74, 31.46), but given the wide confidence intervals, estimates should be interpreted cautiously. The alternative specifications of the event model showed no substantive changes to our inferences, though notably the model extending the event period to 12 weeks showed continued significant associations with the policy at 12 weeks, again with wide confidence intervals.

**Appendix Figure D5** examines trends in incidence in treatment cities when they actually reopened indoor dining. The figure displays trends in 7-day average case rates before and after opening indoor dining and clearly shows an increase in cases beginning about 14 days after reopening, in both groups. **Appendix Figure D6** shows that pre-treatment death rates for treatment and comparison groups do not appear to be parallel, suggesting substantive differences in trends between the groups. We do not provide direct estimates of the association between indoor dining and deaths because of likely differences in the trends, even in absence of the treatment.

## Discussion

In this study of 11 large cities in the US with a total of over twenty-two million inhabitants, we found that keeping indoor dining closed significantly reduced COVID-19 incidence as compared to re-opening. We approached this question leveraging the heterogeneity in state and local policies, specifically comparing cities that kept indoor dining closed to cities that were preempted from doing so but would have kept indoor dining closed if allowed to. Our sensitivity analyses all agreed in the direction of association, enforcing the robustness of these findings. We estimate that, after adjusting for other NPIs, COVID-19 rates decreased by 43% over four weeks in cities that kept indoor dining open compared to cities that re-opened.

In a review of 20 articles using different methods, Bilal et al. found that reopening hospitality venues (bars, restaurants, and night clubs) posed a high-risk for increases in COVID-19 incidence, that closing them was among the most effective COVID-19 mitigation strategies, and that these venues were frequent places for superspreading^22^. However, most reviewed studies did not differentiate between indoor and outdoor dining, so could not isolate impacts of indoor dining on COVID-19 rates. For example, a recent CDC quasi-experimental analysis found that re-opening “on-premise” dining was associated with an increase in cases 41-100 days after allowing indoor dining^11^, and another CDC analysis found that people who tested positive were twice as likely to have reported eating at a restaurant^23^. Another US study using fine-grained mobility data found that compared with reopening other non-residential locations, reopening full-service restaurants was associated with the largest predicted impact on infections^1^. A quasi-experimental study found that closures of restaurant dining areas, in addition to entertainment venues, gyms, and bars, had impacts similar in magnitude to shelter-in-place orders^4^. And, a contact tracing study from Hong Kong found that social venues, including restaurants, accounted for 33% of all traced transmissions, and present an elevated risk for large outbreaks^24^.

While research consistently suggests increases in COVID-19 due to indoor dining, our article is the first to isolate indoor dining independent of outdoor dining using a quasi-experimental design. Additionally, our comparison group satisfies several conditions suggesting a strong counterfactual and our use of preemption improves upon prior analysis that cannot control for differences influencing the likelihood that a geography implements an NPI (selection into the comparison group). The analysis also highlights differences in state and local policy decision powers and authorities, a key policy pandemic question as local government often enacted, or attempted to enact, stricter NPI policies than state governors, producing heterogeneous within-state COVID-19 patterns^25^. Indoor dining has not only been associated with increased COVID-19 incidence, but also has been shown to potentially be responsible for some of the stark racial/ethnic disparities in COVID-19 incidence and mortality^1^. Higher death rates in Black and Hispanic populations are driven by higher rates of infection and exposure, in large part thought to be due to higher rates of occupational exposure^26^. Analyses of occupation and COVID-19 deaths found that food preparation and serving workers^27^, and food and agricultural workers^28^, had among the highest rates of COVID-19 deaths of all workers, and among food preparation and service workers, Hispanic workers had 8 times higher death rates as white workers. Given disproportionate representation among restaurant workers - Hispanic/Latinos account for 27% of food and restaurant workers but only 17% of the population^29^ - reopening may increase risk of exposure, particularly among Hispanic/Latino populations. While our study could not estimate impacts of indoor dining on inequities in COVID-19, due to lack of longitudinal data on neighborhood level or race specific COVID-19 rates across cities, future studies should extend this work to examine disparate impacts of indoor dining policies.

Restaurant groups and lobbyists sent letters to governors and mayors advocating for restaurant re-opening, and in some cases sued (successfully and unsuccessfully) governors and mayors^30^, citing the lack of research on the relationship between indoor dining and COVID rates and superspreading events^31^. Our study cannot speak to impacts of indoor dining on superspreading events, but we do provide robust evidence that indoor dining bans are effective mitigation measures, evidence that can help US and other country courts make determinations about the public health purpose of indoor dining bans. Nearly all cities and states across the US have re-opened indoor dining in the Spring, despite increasing cases in the Northeast and Michigan. These re-opening decisions involve difficult tradeoffs between protecting public health and preventing further layoffs and restaurant closures but have been made more difficult because of limited strong evidence on the association between indoor dining and COVID-19 spread.

Our study has several limitations. The cities are similar on multiple potential confounders that may affect COVID incidence and NPI implementation and compliance, including political leaning, age structure, socioeconomic and housing factors, service workers percentage and mask compliance. However, only a small number of cities met study inclusion criteria, and treatment cities were smaller, had larger, Black and smaller Hispanic populations, and used transit more frequently than comparison cities. These differences should not bias our difference-in-difference analysis unless they were changing significantly during the study period. However, our treatment and comparison groups may differ on other factors related to COVID-19 rates, and our approach does not capture relevant unmeasured or unobservable time-varying between-city confounders, that may have increased COVID-19 transmission, such as mobility data or longitudinal test count, which is not available at the city/county level. Re-opening indoor dining timing in some states coincided with other re-openings such as museums, malls, and theaters. Though contact tracing data suggests that such activities pose relatively limited risk for COVID-19 transmission^32^, if re-opening other non-essential leisure activities also contributed to increased infections then our results may slightly overestimate the association attributable to indoor dining. Importantly, and given transmission dynamics of an infectious disease like COVID-19, the difference between groups in pre-trend levels of COVID-19 incidence may impact the subsequent rate of change^21^, and people may have been less cautious in cities with lower baseline rates. Our model measures associations with indoor dining policies, not the act of engaging in indoor dining specifically, so also captures cases due to policy-related behavior change, and policies re-opening dining may signal reduced risk to residents, leading to more risky activities. Last, we only measured a binary indicator of re-opening (open/closed), though different capacity levels (e.g., 50% vs 25%) or restrictions may differentially impact COVID spread^1^ and future research should examine the impacts of these forms of policy implementation.

## Public Health Implications

Evidence from the 1918 influenza pandemic showed that cities with more stringent public health measures had better economic performance following the pandemic^33^. The closure of indoor dining has devastated the restaurant industry and employees^31^. Providing economic support to the restaurant industry while indoor dining remains closed, while continuing to allow lower-risk activities such as take-out, delivery, and outdoor dining, can achieve COVID-19 mitigation goals without sacrificing economic security. Moreover, if we hope to re-open or keep open schools, a priority for the Biden administration^34^, COVID-19 incidence rates should be as low as possible^35^. Our study shows that one way to achieve this is through keeping indoor dining closed.

Keeping indoor dining closed is one tool to prevent further spread of COVID-19. Cities and states have re-opened indoor dining in the spring, despite increasing cases on the East Coast and Michigan, and increasing predominance of more contagious strains. Though vaccination rates are rapidly increasing in the US, large vulnerable populations remain unvaccinated, and other countries have far lower vaccination coverage. This study suggests that keeping indoor dining closed can prevent thousands of COVID-19 cases.

## Data Availability

Data is from the BCHC COVID-19 Health Inequities in Cities dashboard and from the Center for Systems Science and Engineering (CSSE) at Johns Hopkins University publicly available from the Center for Systems Science and Engineering (CSSE) at Johns Hopkins University.

https://github.com/CSSEGISandData/COVID-19

## Acknowledgements

We would like to acknowledge Dr. Kathryn M. Leifheit for methodological assistance and Dr. Noah Weber for providing a methodological review.

