## Appendix for "Evaluating the impact of keeping indoor dining closed on COVID-19 rates among large US cities: a quasi-experimental design"

### **Appendix A: Dates of Re-openings and Non-Pharmaceutical Interventions**

#### Appendix Figure A1. City and State Re-openings and Rolling 7-Day Average Case Rates

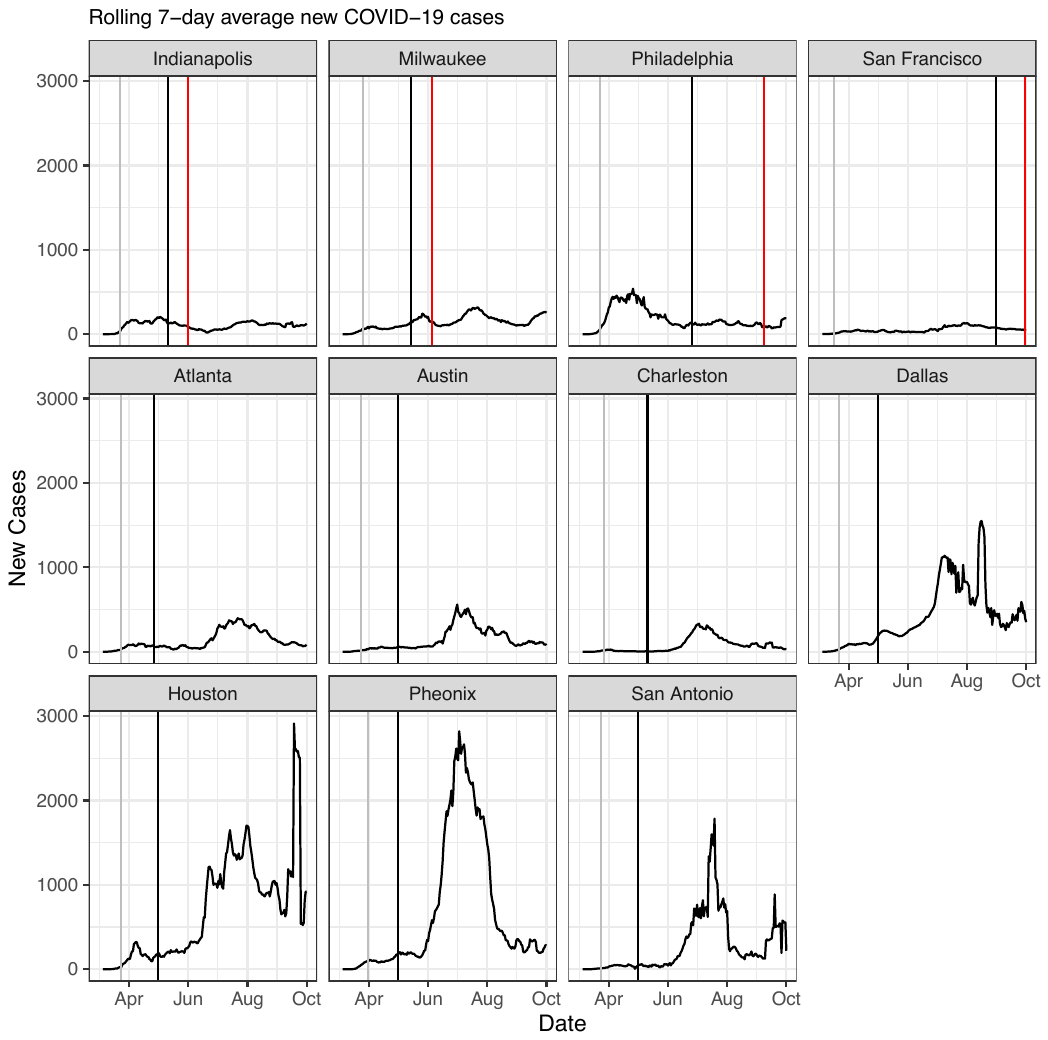

Grey vertical line indicates date when cities closed dining, black vertical line indicates date when state allowed reopening or required re-opening (in comparison cities), red vertical line indicates actual city re-opening in treatment cities (i.e. cities that were allowed to keep indoor dining closed).

#### Appendix Table A2. City and State Re-opening Indoor Dining Dates

| **City and State NPIs-Reopening Indoor Dining and Resuming In-Person Classes** | | | | | |
| --- | --- | --- | --- | --- | --- |
| **Group** | **State** | **City** | **County** | **Indoor Dining Reopen Date-State/Allowed by State** | **Indoor Dining Reopen Date-City** |
| Treatment | California | San Francisco* | San Francisco | 8/31/2020 | 9/30/2020 |
| Treatment | Indiana | Indianapolis* | Marion | 5/11/2020 | 6/1/2020 |
| Treatment | Pennsylvania | Philadelphia* | Philadelphia | 6/26/2020 | 9/8/2020 |
| Treatment | Wisconsin | Milwaukee* | Milwaukee | 5/14/2020 | 6/5/2020 |
| Comparison | Arizona | Phoenix | Maricopa | 5/11/2020 | 5/11/2020 |
| Comparison | Georgia | Atlanta | Fulton | 4/27/2020 | 4/27/2020 |
| Comparison | South Carolina | Charleston* | Charleston | 5/11/2020 | 5/11/2020 |
| Comparison | Texas | Austin | Travis | 5/1/2020 | 5/1/2020 |
| Comparison | Texas | Dallas | Dallas | 5/1/2020 | 5/1/2020 |
| Comparison | Texas | Houston * | Harris | 5/1/2020 | 5/1/2020 |
| Comparison | Texas | San Antonio* | Bexar | 5/1/2020 | 5/1/2020 |

We collected information on statewide and city/county indoor dining orders by searching multiple publicly available databases, and state/city website listing these orders and reviewing state/city orders issued between April 1 and September 30. Our search includes city, county, or state orders signed by Governors, Mayors, or judges. * indicates cities that share contiguous boundaries with the county

#### Appendix Table A3: Dates of City and State Non-Pharmaceutical Interventions

|  |  | | | **City and State NPIs-Stay-At-Home Orders, Mask Mandates, & Eviction Moratoriums** | | | | | | | |  |
| --- | --- | --- | --- | --- | --- | --- | --- | --- | --- | --- | --- | --- |
| **Group** | | **State** | **City** | | **Date City Closed Indoor Dining** | **Date of State Stay-at-home Order Lifted** | **Date of City Stay-at home Order Lifted** | **Date of State Mask Mandate** | **Date of City Mask Mandate** | **State Eviction Moratorium Dates** | **City Eviction Moratorium Dates** | **Public Schools Resume In-Person Classes-City** |
| Treatment | | California | San Francisco | | 3/17/2020 | 5/14/2020 | 5/14/2020 | 6/18/2020 | 4/17/2020 | 3/1/2020 -6/30/2021 | 3/1/20-  6/30/21 | Did not resume in-person classes |
| Treatment | | Indiana | Indianapolis | | 3/23/2020 | 5/1/2020 | 5/18/2020 | 7/27/2020 | 7/9/2020 | 3/19/2020-8/14/2020 | Same as state | Did not resume in-person classes |
| Treatment | | Pennsylvania | Philadelphia | | 3/23/2020 | 6/5/2020 | 6/5/2020 | 7/1/2020 | N/A | 5/7/2020-8/31/2020 | 3/18/2020-12/31/2020 | Did not resume in-person classes |
| Treatment | | Wisconsin | Milwaukee | | 3/26/2020 | 5/13/2020 | 6/5/20 | 8/1/2020 | 5/14/2020 | 3/27/2020-5/27/2020 | Same as state | Did not resume in-person classes |
| Comparison | | Arizona | Phoenix | | 3/31/2020 | 5/16/2020 | 5/16/20 | No mandate* | 6/20/2020 | 3/24/2020-10/31/2020 | Same as state | Did not resume in-person classes |
| Comparison | | Georgia | Atlanta | | 3/24/2020 | 5/1/2020 | 5/1/20 | No mandate | 7/8/2020 | N/A | 3/16/2020-10/31/2020 | Did not resume in-person classes |
| Comparison | | South Carolina | Charleston | | 3/27/2020 | 5/4/2020 | 5/31/20 | No mandate* | 7/1/2020 | 3/17/2020-5/15/2020 | Same as state | 9/8/2020 |
| Comparison | | Texas | Austin | | 3/24/2020 | 5/1/2020 | 7/15/20 | 7/3/2020 | 6/22/20 | 3/19/2020-5/19/2020 | 3/19/2020-12/31/2020 | 10/5/2020 |
| Comparison | | Texas | Dallas | | 3/22/2020 | 5/1/2020 | 5/15/20 | 7/3/2020 | 6/19/2020 | 3/19/2020-5/19/2020 | Same as state | 9/28/2020 |
| Comparison | | Texas | Houston | | 3/24/2020 | 5/1/2020 | 5/20/20 | 7/3/2020 | 4/22/2020 | 3/19/2020-5/19/2020 | Same as state | 10/19/2020 |
| Comparison | | Texas | San Antonio | | 3/24/2020 | 5/1/2020 | 6/5/20 | 7/3/2020 | 6/17/2020 | 3/19/2020-5/19/2020 | 3/19/2020-6/1/2020 | 9/8/2020 |

We collected information on statewide and city/county orders from searching multiple publicly available databases, and state/city websites listing these orders and reviewing state/city order. *Masks mandated for certain businesses, no broad state-wide mask mandate.

### **Appendix B: Counterfactual Assumption & Re-Opening Citations**

#### Appendix Table B2: Citations for Counterfactual Assumption that Cities Would Have Kept Indoor Dining Closed if Not Preempted, Comparison Cities

| **Evidence of Preemption** | | | |
| --- | --- | --- | --- |
| **City** | **State** | **Preempted City Policy** | **State Preemption** |
| **Phoenix** | **Arizona** | [Mayor Gallego Tweet](https://twitter.com/MayorGallego/status/1281338056058867714)  [Mayor Gallego Statement On Stay-at-Home Order](https://www.phoenix.gov/newsroom/mayors-office/1151) | [Arizona Executive Order](https://azgovernor.gov/sites/default/files/eo_2021_0.pdf) |
| **Austin** | **Texas** | [Interview with Mayor Adler](https://www.nbcnews.com/news/us-news/some-texas-cities-nervous-governor-reopens-state-everybody-scared-n1198691)  [April 13, 2020 Executive Order](https://www.austintexas.gov/sites/default/files/files/document_96DEBEEC-E581-05E0-8A3D444404948A84.pdf)  [May 8, 2020 Executive Order](https://www.austintexas.gov/sites/default/files/files/Order%2020200508-011.pdf) | [Texas Executive Order](https://gov.texas.gov/news/post/governor-abbott-issues-executive-order-relating-to-the-expanded-reopening-of-services) |
| **Dallas** | **Texas** | [Letter to Honorable Clay Jenkins of Dallas County](https://www.texasattorneygeneral.gov/sites/default/files/images/admin/2020/Press/Dallas%20County%20Letter_05122020.pdf?)  [Dallas County April 23, 2020 Executive Order](https://www.dallascounty.org/Assets/uploads/docs/covid-19/orders-media/042320-DallasCountyOrder.pdf)  [Dallas County May 4, 2020 Executive Order](https://www.dallascounty.org/Assets/uploads/docs/covid-19/orders-media/050120-Supplemental-DallasCountyOrderonReopenedServices.pdf) |  |
| **San Antonio** | **Texas** | [Texas City and County Leaders ask Gov Greg Abbott for authority to implement local stay-at-home orders](https://www.texastribune.org/2020/06/29/texas-coronavirus-stay-at-home-harris-dallas/)  [Executive Order NW-05 OF County Judge Nelson W. Wolff, Issued April 6, 2020 (bexar.org)](https://www.bexar.org/DocumentCenter/View/26538/Executive-Order-NW-05-April-6-2020?bidId=)  [Executive Order NW-07 OF County Judge Nelson W. Wolff Issued April 29, 2020 (bexar.org)](https://www.bexar.org/DocumentCenter/View/26838/Executive-Order-NW-07-Issues-April-29?bidId=) |  |
| **Houston** | **Texas** | [Mayor Turner's letter to Governor Abbott](https://www.chron.com/file/650/7/6507-6.30.2020%20MST%20to%20Abbott.pdf)  [Harris County April 4, 2020 Executive Order](https://agenda.harriscountytx.gov/2020/AmendedStayHomeOrder.pdf)  [Harris County May 1, 2020 Executive Order](https://agenda.harriscountytx.go/)  v/2020/20200501Order.pdf |  |
| **Atlanta** | **Georgia** | [Atlanta Isn't Ready to Reopen-And Neither is Georgia-Keisha Lance Bottoms in The Atlantic](https://www.theatlantic.com/ideas/archive/2020/04/its-too-early-to-reopen-georgia/610909/) | [2020 Executive Orders \| Governor Brian P. Kemp Office of the Governor (georgia.gov)](https://gov.georgia.gov/executive-action/executive-orders/2020-executive-orders) (See Executive Order 7/15/20 Pg 32) |
| **Charleston** | **South Carolina** | [2020-070-Emergency-Ordinance---Stay-at-Home (charleston-sc.gov)](https://www.charleston-sc.gov/DocumentCenter/View/27264/2020-070-Emergency-Ordinance---Stay-at-Home) | [2020-05-08 FILED Executive Order No. 2020-34 - Authorization of Limited Indoor Dining Services & Rescission of Boating Restrictions.pdf](https://governor.sc.gov/sites/default/files/Documents/Executive-Orders/2020-05-08%20FILED%20Executive%20Order%20No.%202020-34%20-%20Authorization%20of%20Limited%20Indoor%20Dining%20Services%20%26%20Rescission%20of%20Boating%20Restrictions.pdf) |

We collected information on statewide and city/county orders from searching multiple publicly available databases, and state/city websites listing these orders and reviewing state/city orders. We identified public statements by searching news articles, twitter posts, and state/city websites.

#### Appendix Table B3: Information and Evidence for City and State Indoor Dining Reopening Orders, Treatment Cities

| **Opening Date References for Treatment Cities** | | | | |
| --- | --- | --- | --- | --- |
| **City** | **State** | **Detail** | **City Evidence** | **State Evidence** |
| **Indianapolis** | **Indiana** | Governor Holcomb permitted indoor dining to resume throughout Indiana on 5/11/2020. Indianapolis waited until 6/1/2020 to resume indoor dining despite being eligible to reopen indoor dining on 5/11. | [Mayor Joe Hogsett Statement on Reopening Indianapolis](https://citybase-cms-prod.s3.amazonaws.com/455ab1988f7f491684889f6200d1ce1e.pdf) | [Back On Track Indiana: Governor Holcomb's Indiana COVID-19 Update (May 1, 2020)](https://www.backontrack.in.gov/2362.htm) |
| **Philadelphia** | **Pennsylvania** | PA permitted Philadelphia County to resume indoor dining on 6/26/2020 when the city entered the green phase. Philadelphia waited until 7/3/2020 to enter the green phase for most activities, but the city waited until 9/8/2020 to resume indoor dining. | [What the green phase means for Philadelphia \| Department of Public Health \| City of Philadelphia](https://www.phila.gov/2020-06-18-what-the-green-phase-means-for-philadelphia/#:~:text=When%20Philadelphia%20enters%20the%20green,.%20Schools%20and%20colleges.)  [Philadelphia to Reopen Indoor Dining Sept 8, 2020](https://www.phila.gov/2020-08-20-indoor-dining-is-back-on-september-8-in-philadelphia/#:~:text=Indoor%20dining%20is%20back%20in,while%20gradually%20and%20safely%20reopening) | [Gov. Wolf: 12 More Counties to Go Green on June 26 (pa.gov)](https://www.governor.pa.gov/newsroom/gov-wolf-12-more-counties-to-go-green-on-june-26/)  [Process to Reopen Pennsylvania (pa.gov)](https://www.governor.pa.gov/process-to-reopen-pennsylvania/) |
| **San Francisco** | **California** | Governor Newsom issued county guidance preempting counties from enacting less restrictive policies than those directed by the state's mandate. While the city of San Francisco met statewide guidelines (red phase) for re-opening indoor dining on 8/28, the city chose to keep indoor dining closed. On 9/30 the city issued guidance allowing indoor dining at 25%. | [San Francisco to Move Forward with Reopening More Businesses and Activities on September 30 \| Office of the Mayor (sfmayor.org)](https://sfmayor.org/article/san-francisco-move-forward-reopening-more-businesses-and-activities-september-30) | [CA Phased Re-opening Order](https://www.gov.ca.gov/wp-content/uploads/2020/05/5.4.20-EO-N-60-20-text.pdf%20)  [Dashboard for County Phases, as of August 28, 2020](https://sfist.com/2020/08/28/newsom-new-color-codes-covid-county-watch-list/) |
| **Milwaukee** | **Wisconsin** | Wisconsin’s Safer At Home order went into effect on March 25, 2020, it prohibited nonessential travel and limited gatherings.  On April 20th the governor issued the "Badger Bounce Back" plan to continue the emergency order and determine phased re-opening plans. On May 13th the Wisconsin state supreme court struck down the executive order. On May 14th Milwaukee issued their own order, and though allowed by the state to open up as of 5/14, did not re-open indoor dining until 6/5. | [Milwaukee May 14 Order](https://www.documentcloud.org/documents/6890535-Moving-Milwaukee-Forward.html)  [Milwaukee Phase 3 Order](https://city.milwaukee.gov/MMFSReleasePh3)  [Milwaukee Phase 4 Order](https://city.milwaukee.gov/ImageLibrary/MKE-Health1/MMFSReleasePh4_6.26.20.pdf) | [Wisconsin Safer at Home Order](https://content.govdelivery.com/accounts/WIGOV/bulletins/282deef)  [Wisconsin Supreme Court Strikes Down Stay-at-Home Order](https://www.nytimes.com/2020/05/13/us/coronavirus-wisconsin-supreme-court.html) |

We collected information on statewide and city/county orders from searching multiple publicly available databases, and state/city websites listing these orders and reviewing state/city orders. We identified public statements by searching news articles, twitter posts, and state/city websites.

### **Appendix C: Model Specification**

#### Appendix C1: Model Specification and Description

The main model specification is as follows:

$${log(daily cases)}_{ik} =\beta_{0}+\beta_{1}{Post}_{ik}+\beta_{2}Dining Closed_{k}+\beta_{3}{DiningClosed_{k}*Post}_{ik}+\beta_{4}X_{ik}+{ln(population)}_{k}$$

Where we model the daily count of cases in each *k*th city on the *i*th day. For the treatment group the dummy variable *Post* is coded as 1 for days beginning 14 days post time-zero (with time-zero being the day the state reopened but the city remained closed), and 0 otherwise. For the comparison group, the variable Post is coded as 1 for days beginning 14 days post time-zero (with time zero being the date the city re-opened indoor dining because it was preempted by the state), and 0 otherwise. The *Dining Closed* dummy variable is 1 in treatment cities (that kept indoor dining closed) and 0 in comparison cities (that attempted to keep indoor dining from opening but were preempted from doing so). Our main coefficient of interest is $\beta_{3}$: by exponentiating $\beta_{3}$ we obtain the incidence rate ratio representing the association between keeping indoor dining closed compared with re-opening indoor dining. $\beta_{4}X_{ik}$ represents other time-varying local mitigation & reopening policies that may act as potential confounders: stay at home orders, mask mandates, and city/state eviction bans, with a 14-day lag from the date of policy implementation. We use a negative binomial model, with city population as the offset and with robust standard errors clustered at the city level.

### **Appendix D: Robustness Checks**

#### Appendix D1: Description of sensitivity analysis

As detailed in the main manuscript, we conducted six sets of sensitivity analyses to test for robustness of our model to alternative specifications: we: (1) extended the duration of follow-up to 12 weeks and varied the pre-specified 2-week lag, (2) excluded various cities, (3) included calendar fixed-effects, (4) used the dates treatment cities actually re-opened indoor dining rather than the dates the state allowed the treatment cities to re-open, (5) used an event study model, an alternative specification of the DiD model, and (6) repeated the analysis using death rates instead of case rates.

First, the main analysis includes a 6-week (2-week lag + 4-weeks post lag) follow-up period, and we tested extending the follow-up period to 12 weeks. Our main analysis also uses a 2-week lag to account for delays from infection to case reporting, and we considered alternative lag periods of 9-days, 3-weeks, and 4-weeks. Second, we limited our analysis to only the cities in the BCHC, to test if city selection impacts estimated impacts. We additionally tested removing San Francisco (state allowed the city to open 8/31) from the analysis, to test if patterns of declined cases after delaying re-opening were explained by seasonal differences. Third, to control for any time varying between city seasonal or disease dynamic effects we also included a calendar week fixed effect in the main model. Fourth, to further assesses if changes in case rates appeared to be related to re-opening indoor dining, we compared case rates in treatment cities before and after the cities actually reopened, using the city-specific dates of re-opening rather than the dates the state allowed the treatment cities to re-open (dates in the column, “Indoor Dining Reopen Date-City” in Appendix Exhibit A1), but do not conduct any formal tests.

Fifth, we employ an event study specification, which is similar to our main difference-in-differences model, to examine whether the effects of re-opening indoor dining differ in the weeks following re-opening. The event-study specification has been used recently to examine the effect of eviction moratoriums and mask mandates, and on-premise dining on COVID-19 incidence^1-3^. We estimated the impact of re-opening, rather than keeping indoor dining closed as the treatment, because we expected a change in trend after re-opening. Though the model allows for estimation of changing growth rates by week, it does not take advantage of our counterfactual comparison group, because it uses the date of re-opening for all cities, rather than comparing cities that re-opened to those that remained closed, so may be subject to additional selection bias.

We estimate the following model, using city-level daily data:

$${log(daily cases)}_{ik} =\beta_{0}+\beta_{1}{Pre\_week4}_{ik}+\beta_{2}{Pre\_week3}_{ik}+ \beta_{3}{Pre\_week2}_{ik}+\beta_{4}{Post\_week1}_{ik} +\beta_{5}{Post\_week2}_{ik}+\beta_{6}{Post\_week3}_{ik}+\beta_{7}{Post\_week4}_{ik}+\beta_{8}{Post\_week5}_{ik}+\beta_{9}{Post\_week6}_{ik}+ \beta_{10}{Post\_week7}_{ik}+\beta_{11}{Post\_week8}_{ik}+ \beta_{12}X_{ik}+{ln(population)}_{k}$$

We measured associations between the re-opening and COVID-19 case rates using a reference period (1 week before re-opening ^1^) compared with 11 mutually exclusive weeks relative to implementation. The coefficients on the post weeks represent the impact of re-opening indoor dining during each of those post re-opening weeks, compared to the week before re-opening. We expect the associations to grow over time if there is increasing use of indoor dining and if indoor dining increases infection risk. The pre-week variables capture differences in pre-reopening COVID-19 trends before cities re-opened indoor dining.

To control for the time-varying impacts of other NPIs, we added $\beta_{12}X_{ik}$ to the model, representing other time-varying local mitigation & reopening policies that may act as potential confounders: stay-at-home orders, mask mandates, and city/state eviction bans, with a 14-day lag from date of policy implementation. These are included as ordinal variables (instead of binary ones), using the count of weeks since the NPI was implemented (1-5, with >4 weeks after implementation coded as 5). If a city never enacted the policy, or enacted the policy outside of the study period, we code the variable as zero for all observations. Finally, we add a control for calendar date, to control for any secular time trends in the mean outcome^4^, and applied an unconditional fixed effect negative binomial approach using city dummies^5^ to control for time-invariant between city factors. We tested extending the study period to 12 weeks after re-opening (alternative specification 1), and to setting the NPI confouders as ordinal variations without the 4+ week limitation (i.e. NPI ordinal variables can go above 5 after 5 weeks post implementation; alternative specification 2).

Sixth and last, we repeated our main analysis but included death rates as our outcome rather than case rates. We considered a main analysis with a 5-week lag (2-week case + 3-week lag after cases) and a 6-week lag. We limit this analysis to the 2 weeks before and 9-weeks after re-opening. Pre-period trends did not appear parallel for deaths, so we only present descriptive mortality results for this analysis.

#### Appendix Table D2: Robustness Checks: Alternative lags, periods, cities, and calendar week fixed effects

| **Incidence Rate Ratios** | |
| --- | --- |
| *Main Model (adjusted)* | 0.570^***^ (0.37, 0.87) |
| *Sensitivity Model 5*: Extend follow-up to 12 weeks | 0.178^***^(0.11, 0.28) |
| *Sensitivity Model 1*: no lag for other NPIs | 0.556^**^ (0.33, 0.95) |
| *Sensitivity Model 2*: 9-day lag | 0.681^**^ (0.46, 0.99) |
| *Sensitivity Model 3:* increase lag to 3 weeks (3 weeks) | 0.416^***^ (0.24, 0.72) |
| *Sensitivity Model 4*: increase lag to 4 weeks (28 days) | 0.291^***^ (0.16, 0.52) |
| *Sensitivity Model 6*: remove the non BCHC cities | 0.525^***^ (0.34, 0.8) |
| *Sensitivity Model 7:* Remove CA (Early fall re-opening) | 0.440^***^ (0.26, 0.75) |
| *Sensitivity Model 8:* Adding Calendar week fixed effects | 0.430^**^ (0.22, 0.93) |

Results come from a negative binomial model with city population model as an offset, robust standard errors clustered at the city level, and a two-week case lag. Models adjusted for mask mandates, stay at home orders, and eviction moratoriums. *p<0.1; **p<0.05; ***p<0.01

#### Appendix Table D3: Event Study Adjusted Incidence Rate Ratios

|  |  | **Alternative Specifications** | |
| --- | --- | --- | --- |
|  | **Main Model** | **1** | **2** |
| **Weeks since lifted** | **IRR(CI)** | **IRR(CI)** | **IRR(CI)** |
| -4 | 0.59 (0.81, 1.23) | 0.61 (0.33, 1.11) | 0.53 (0.27, 1.02) |
| -3 | 0.81 (0.5, 1.24) | 0.84 (0.5, 1.41) | 0.78 (0.44, 1.37) |
| -2 | 1.00 (0.34, 1.0) | 0.97 (0.78, 1.22) | 0.97 (0.79, 1.18) |
| -1 | Ref | Ref | Ref |
| 1 | 1.18 (0.82, 1.69) | 1.17 (0.82, 1.66) | 1.47 (0.94, 2.32) |
| 2 | 1.25 (0.66, 2.35) | 1.19 (0.64, 2.22) | 1.55 (0.74, 3.23) |
| 3 | 1.30 (0.53, 3.23) | 1.12 (0.47, 2.7) | 1.84 (0.84, 4.00) |
| 4 | 1.73 (0.55, 5.43) | 1.38 (0.42, 4.56) | 2.31 (0.88, 6.09) |
| 5 | 3.52^**^ (1.16, 10.69) | 2.75 (0.85, 8.83) | 3.38^**^ (1.23, 9.29) |
| 6 | 5.49^***^ (1.82, 16.56) | 4.34^**^ (1.4, 13.52) | 4.62^***^ (1.7, 12.5) |
| 7 | 9.30^***^ (2.98, 29.05) | 7.04^***^ (2.16, 22.99) | 6.6^***^ (2.32, 18.77) |
| 8 | 10.85^***^ (3.74, 31.46) | 9.91^***^ (3.61, 27.18) | 10.66^***^ (4.93, 23.0) |
| 9 |  | 9.18^***^ (4.43, 19.03) | 13.03^***^ (8.72, 19.48) |
| 10 |  | 9.39^**^ (4.57, 19.31) | 16.18^***^ (10.46, 25.03) |
| 11 |  | 7.40^**^ (3.43, 15.97) | 15.16^***^ (8.57, 26.82) |
| 12 |  | 6.21^**^ (2.78, 13.8) | 12.93^***^ (7.01, 23.86) |

Results come from a negative binomial model with city population model as an offset, the week before re-opening as reference, and robust standard errors clustered at the city level. Models further adjusted for mask mandates, stay at home orders, eviction moratoriums, and week and city fixed effects. *Alternative Specification 1*: period extended to 10 weeks pre and 12 post (assumed 2-week lag). *Alternative Specification 2:* specification 1 + no NPI 4+ week limitation. *p<0.1; **p<0.05; ***p<0.01

#### Appendix Figure D4: Event Model

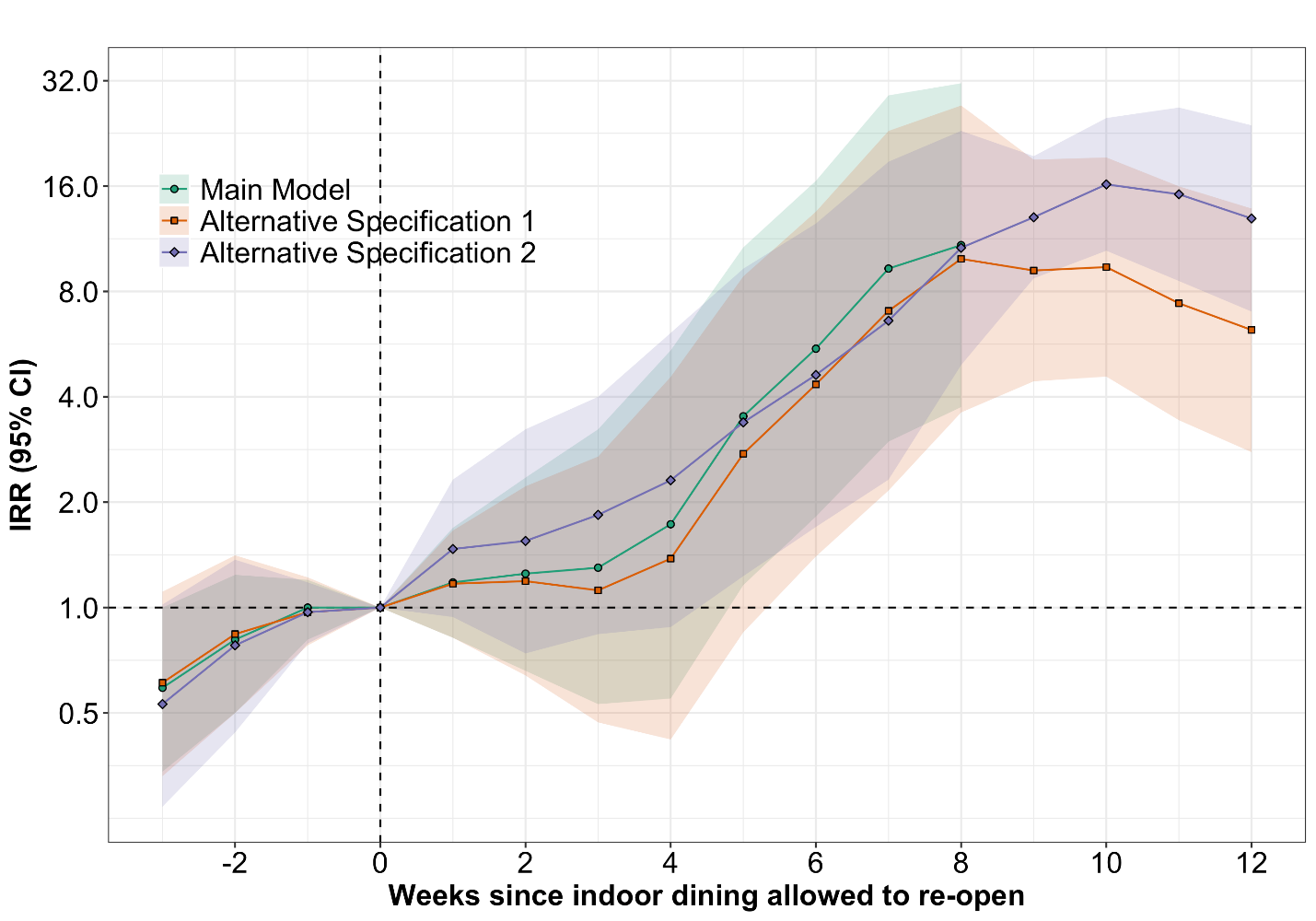

#### Appendix Figure D5: Trends in Case Rates Comparing Treatment and Comparison Groups, Using Date of Actual City Re-opening

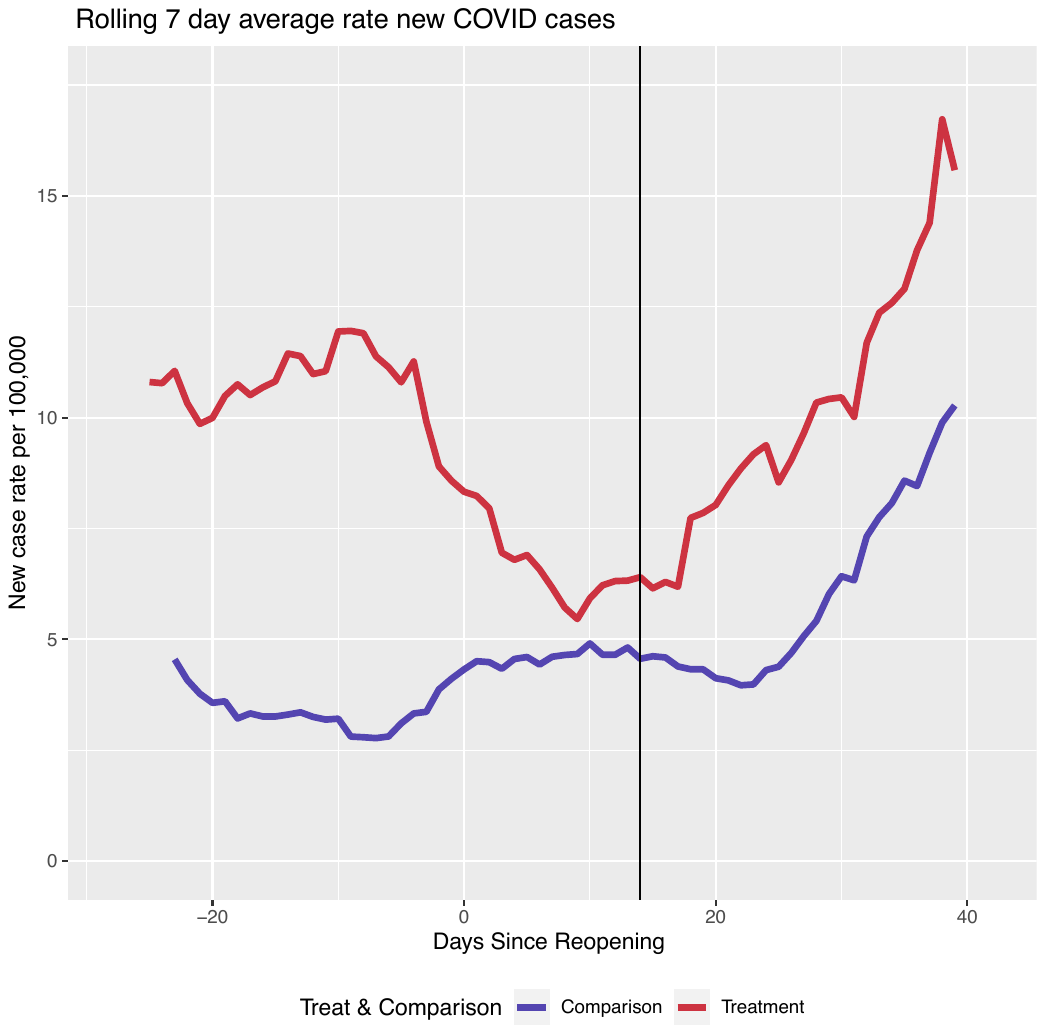

Comparison cities include Atlanta, Austin, Charleston, Dallas, Houston, Phoenix, and San Antonio; treatment cities include Indianapolis, Milwaukee, Philadelphia, and San Francisco. Vertical black line represents a 2-week lag after re-opening in both treatment and comparison cities.

#### Appendix Figure D6: Trends in Death Rates Comparing Treatment and Comparison Groups

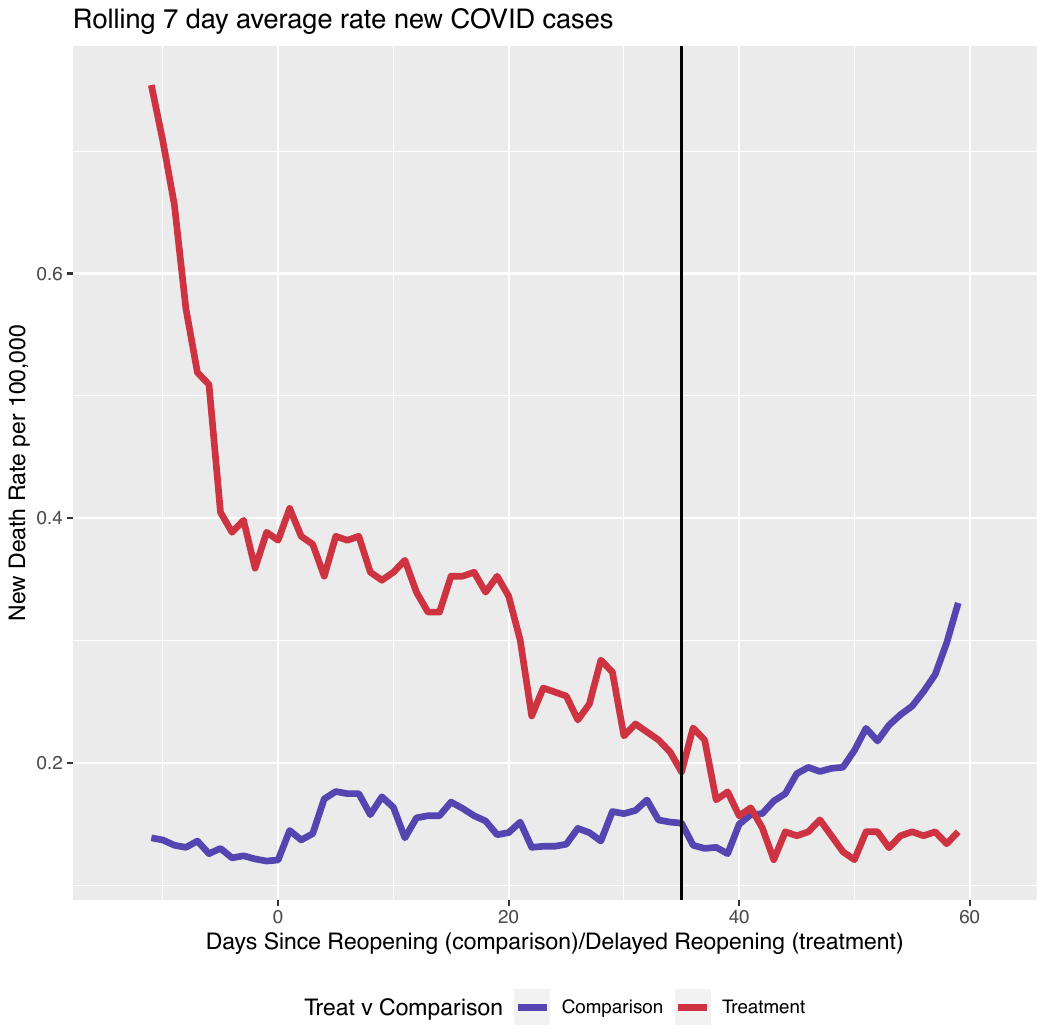

SOURCE: Authors’ analysis of city/county-level COVID-19 death data between April and November of 2020. NOTES: comparison cities include Atlanta, Austin, Charleston, Dallas, Houston, Phoenix, and San Antonio; treatment cities include Indianapolis, Milwaukee, Philadelphia, and San Francisco. Vertical black line represents a 35-day lag after re-opening (comparison cities) or the date the state allowed the city to re-open, but the city stayed closed (treatment cities).
